# Automated Detection of Speech Disorders in Parkinson’s Disease using Deep Convolutional Neural Networks: A Pilot Study

**DOI:** 10.1101/2025.07.18.25331737

**Authors:** Sara A. Jones, Jeremy Cosgrove, He Wang, Ryan K. Mathew

## Abstract

**Background:** Patients with Parkinson’s disease (PD) frequently exhibit deficits in functional communication due to the presence of speech disorders associated with dysarthria that can be characterized by monotony of pitch (or fundamental frequency), reduced loudness, irregular rate of speech, imprecise consonants, and changes in voice quality. This pilot study investigates the application of a speech classifier based on deep-convolutional neural networks (CNNs) for aiding early diagnosis of PD.

**Methods:** In this study, we analyse the performance capabilities of two audio feature extraction techniques and associated model architectures: low-level time-frequency based features classified using a Support Vector Machine (SVM); and classifying log mel-spectrograms of segmented audio signals using varying depths of Deep Convolutional Neural Networks. The models were trained using an open-source data set comprised of 73 audio recordings of continuous dialogue from 37 subjects, including 16 people with PD (5 females and 11 males) and 21 healthy controls (19 females and 2 males), who were required to perform two speech production tasks.

**Results:** The experimental results show that the deep CNN model, trained on the log mel-spectrograms of 5-second segmented audio signals, can successfully differentiate PD subjects from healthy controls (HC) with a mean accuracy of 84.7%, sensitivity of 87.9% sensitivity and specificity of 89.4%, thus demonstrating its potential for aiding early diagnosis of PD in a clinical setting. The saliency maps show that the deep CNN model can distinguish between PD participants and healthy controls by detecting centralised, low-frequency regions of the spectrograms representing the speech of PD subjects, whereas a larger range of frequencies are detected in the spectrograms representing speech from healthy controls.

## 1 Introduction

Parkinson’s disease (PD) is a chronic, progressive, incurable neurodegenerative disorder resulting from the degeneration and subsequent loss of dopaminergic neurons within the substantia nigra, part of the basal ganglia within the brain. The disease, which affects approximately 0.2*%* (incidence of 15-20 per 100,000) of the general population in the UK [1], is currently diagnosed based on motor parkinsonism - the presence of bradykinesia (slowness of movement) in combination with either rigidity or resting tremor [2]. However, approximately 50–70*%* of nigrostriatal dopaminergic neurons have degenerated before neurologists and geriatricians are able to establish the diagnosis of PD according to this widely accepted clinical diagnostic criteria [3]. There is substantial research attention on the early detection of PD, as this would allow more focused clinical trials and earlier delivery of a disease modifying medication, if and when this becomes available. Prior studies indicate that between 70*%* to 90*%* [4][5] of individuals with PD exhibit hypokinetic dysarthria (HD), a motor speech disorder characterised by a reduction in speech intensity (hypophonia), altered voice quality (dysphonia), flattened pitch inflection (monopitch), loss of stress (monoloudness), festination, breathiness, irregular pauses and hesitancy of speech [6]. Evidence suggests that voice dysfunction is the earliest indicator of motor impairment and may manifest 6-7 years before a clinical diagnosis of PD; i.e., preceding the onset of other motor features [7]. Therefore, automated acoustic analysis is considered by many researchers as an effective, non-invasive tool for PD screening and early detection.

Current approaches to detecting neurodegenerative disorders through speech analysis predominately focus on extracting perturbed features from repeated phonations of sustained vowels [8][9][10] to minimise acoustic variability between participants (e.g. lexical variations) and facilitate robust feature extraction. Whilst sustained phonation may be beneficial in eliciting some aspects of dysphonia, recent studies [11][12] suggest that they lack the high-level prosodic information (e.g. variations of fundamental frequency, pauses and phonation onsets) required to detect the wide range of articulatory deficits related to hypokinetic dysarthria. Pompili et al. [11] propose that a more articulatory demanding speech task than sustained phonation is required to capture subtle changes of speech due to neurodegeneration and therefore extract features obtained from natural voice production (running speech) in which subjects are instructed to speak a pre-devised sentence or passage containing representative linguistic units. The work presented here focuses on two complex speech production tasks, spontaneous dialogue and text-dependent running speech, which require sustained attention and therefore have a tendency to elicit articulatory deficits in the speech of PD subjects compared with simple tasks (e.g. repetition of isolated words and vowels). To the best of our knowledge, there are no studies that specifically focus on comparing complex speech production tasks in terms of their utility for automatic PD discrimination.

Several studies have utilised time-frequency based features extracted from speech signals to classify between PD and control subjects, including jitter (temporal perturbation of the fundamental frequency), shimmer (temporal perturbation of the amplitude of the signal), pitch, amplitude perturbation quotient (APQ), noise-to-harmonic ratios (NHR), proportion of the vocalic duration and degree of voiced breaks [13][14]. Whilst short-term fluctuations in periodic vocal samples of phonatory signals can be quantified using low-level acoustic descriptors (LLD) such as jitter and shimmer, they may be unsuitable for severely disordered voices, which are typically characterised by poor periodicity and from which a period of sustained phonation is harder to extract [15].

In recent years, as an alternative to using classical feature extraction methods to model pathological speech, deep learning methods have been successfully implemented to evaluate specific phenomena in speech, including the detection and monitoring of PD speech disorders [16][17]. Korzekwa et al.[17] proposed a novel deep learning approach for the detection and reconstruction of dysarthric speech in which the authors use a Recurrent Convolutional Neural Network (RCNN) to encode the input mel-spectrogram, reconstruct the spectrogram from a low-dimensional dysarthric latent space and encoded text and apply a logistic classification model to predict the probability of dysarthric speech. By contrast, a study into the ability of Convolutional Neural Networks (CNN) to learn representative features directly from raw audio signals reveals that whilst the networks are able to autonomously discover frequency decompositions from raw audio, this method does not outperform spectrogram-based approaches [18]. Piczak [19] demonstrated that by representing an audio input using time-frequency representations (i.e., log-scaled mel-spectrograms), it is possible to directly utilise architectures that were initially developed for image data processing, such as CNNs, to classify environmental and urban audio samples. However, a 2-layered network is considered shallow in comparison to networks used by other applications, which is a shortcoming of the architecture used by [19]. The ability of convolutional networks to generate highly accurate results is dependent on the depth of the network, hence using deeper networks could improve the accuracy of vocal disorder detection. Therefore we intend to demonstrate the capabilities of deep learning in the detection of vocal disorders when deeper networks are used and demonstrate that an increase in the number of layers leads to an increase in the ability of the network in discriminating between PD and control subjects.

This study focuses on identifying the optimal audio feature extraction technique and model architecture for PD vocal disorder detection in spontaneous dialogue and text-dependent running speech. Our contributions can hence be summarized as follows:

- We compare two complex speech production tasks (spontaneous dialogue and text-dependent running speech) in terms of their utility for automatic PD discrimination.
- We analyse the performance capabilities of two audio feature extraction techniques and associated model architectures: low-level time-frequency based features classified using a Support Vector Machine (SVM); and classifying mel-spectrograms of segmented audio signals using varying depths of Deep Convolutional Neural Networks (DCNNs).
- We study how the depth (the number of layers) of the deep neural networks effects the performance of PD vocal disorder detection capability of the network.
- We evaluate and compare the saliency maps generated by the Deep Convolutional Neural Network in identifying regions of the time-frequency waveform that provide an indication of hypokinetic dysarthria.

## 2 Methods

The Mobile Device Voice Recordings at King’s College London (MDVR-KCL) dataset used in this paper is open-source [20], therefore a separate ethics review was not required for the use of open-source data in secondary data analysis. The original study was reviewed and approved by the London - Dulwich Research Ethics Committee (REC reference 17/LO/0909). All subjects signed written informed consent to participate in this study.

### 2.1 Dataset Description and Audio Pre-processing

The Mobile Device Voice Recordings at King’s College London (MDVR-KCL) data set [20] consists of 73 audio recordings of continuous dialogue from 37 subjects, including 16 people with PD (5 females and 11 males) and 21 healthy controls (HC) (19 females and 2 males). Subjects were required to perform two speech production tasks; to recite a passage from the fable “The North Wind and the Sun” and to engage in spontaneous dialog with the test executor about places of interest, local traffic, or personal interests. The participants were interviewed in a standard examination room, measuring approximately 10 × 10 m and with a reverberation time of 0.5 seconds.

The audio signals were recorded using a Motorola Moto G4 Smartphone at a sampling rate of 44.1 kHz with 16-bit resolution. Subjects held the recording device adjacent to their preferred ear, such that the in-built microphone was in direct proximity to their mouth, therefore one can assume that all recordings were performed within the reverberation radius and that the recordings can be considered as “clean”. The length of the recordings vary from 73 to 221 seconds, with an average duration of 143 seconds. The audio samples were assessed by expert neurologists and the speech intelligibility of each participant was scored according to the speech components of the Unified Parkinson’s Disease Rating Scale (UPDRS)-II (Part 5), UPDRS-III (Part 18) and Hoehn & Yahr, which are five-point standard scales that measure the severity and symptom progression of PD. The clinical and demographic characteristics of the participants are presented in Table 1.

**Table 1.** Demographic and clinical characteristics of subjects. *Note*: *n* = number of participants, *PD* = Parkinson’s Disease, *M* = Male, *F* = Female, *H-Y* = Hoehn & Yahr, *UDPRS* = Unified Parkinson’s Disease Rating Scale. Data is expressed as mean ± standard deviation

|  | PD Group (n = 16) | Control Group (n = 21) |
| --- | --- | --- |
| Gender Ratio (M/F) | 11/5 | 2/19 |
| H-Y | $2.63 \pm 0.72$ | N/A |
| UPDRS II-5 | $0.81 \pm 0.91$ | N/A |
| UPDRS III-18 | $0.9375 \pm 0.93$ | N/A |

During pre-processing, we manually removed all audio segments containing dialogue spoken by the test executor, which may have otherwise negatively affected the performance of the classifiers in distinguishing between PD and HC participants. In order to augment the size of the data set, we employed a sliding window approach [21] [22] [23] to divide the original audio signal into 5 and 10 second segments with a 50% overlap in the temporal dimension between two consecutive segments, thereby generating 1575 five second segments and 806 ten second segments. Since any element of the audio signal is contained within two consecutive segments, with the exception of the first and last segments, any information loss at a boundary of a segment is recovered by referencing to the adjacent overlapping segment. The length of segment was chosen as this duration was deemed sufficient to capture slow-changing, long-range temporal features of the speech waveform.

### 2.2 Feature Extraction

#### 2.2.1 Low-Level Acoustic Features

We extracted 24 low-level quantitative vocal parameters from each audio segment using Parselmouth [24], a Python library which provides a wide range of algorithms related to the acoustic analysis, manipulation, and synthesis of speech signals. To assess the auditory features of vocal dysfunction in PD, we examined parameters relating to: rate of vocal fold vibration (fundamental frequency, F_0_, or pitch), frequency perturbation (jitter), amplitude perturbation (shimmer), changes in vocal quality (harmonics-to-noise ratio, HNR), and aperiodicity (degree of voicelessness, DUV), amongst others. Functionals characterizing statistical and temporal properties of these low-level descriptors were also computed. Linguistic features have not been extracted since the purpose of this study was to analyse the auditory characteristics of vocal dysfunction in PD, regardless of the language used.

#### 2.2.2 Mel-Spectrogram Feature Representation

In this approach, input audio signals are represented as spectrograms in which the time and frequency axes are considered as spatial dimensions so that a CNN model, commonly used in image recognition tasks, can be applied. Although any spectrogram variant is applicable, we employed the log mel-scaled spectrogram, as it reduces the resolution of high band frequencies and more accurately resembles the auditory perception capabilities of humans. Utterances shorter than the length of interval (i.e. 5 or 10 seconds) are zero-padded. The mel-scaled spectrogram is extracted as a feature from the audio segments using the Python Librosa library [25] with the original sampling frequency of 44.1 kHz, 1024 Fast Fourier Transform (FFT) points, 128 mel-bins, and a hop-length of 512 frames. The amplitude of the mel-spectrogram is scaled logarithmically, and the scaled mel-spectrogram is resized in the temporal dimension to fit the input size required by the CNN classifier (224 × 224 pixels).

### 2.3 Model Architectures

In this study, we analyse the performance capabilities of two audio feature extraction techniques and associated model architectures: low-level time-frequency based features classified using a Support Vector Machine (SVM); and classifying mel-spectrograms of segmented audio signals using varying depths of Deep Convolutional Neural Networks.

We employed two variants of the densely connected convolutional network (DenseNet) [26] architecture, DenseNet-121 and DenseNet-169, to classify the mel-spectrograms described in Section 2.2.2. The architectures of the DenseNet models are provided in Table 2. DenseNet consists of a series of densely connected convolutional and transitional blocks in which the output feature maps of each layer are concatenated with the output feature maps of all successive layers, thereby providing variety in the inputs of the subsequent layers. Each layer is connected to both the previous layer and all preceding layers in a feed-forward fashion. DenseNets have several compelling advantages: they alleviate the vanishing-gradient problem, strengthen feature propagation and encourage feature re-use, whilst substantially reducing the number of network parameters.

**Table 2.** Sizes of outputs and convolutional kernels for different DenseNet architectures. FC: Fully Connected, conv: convolution. This table has been adapted from [26].

| Layers | Output Size | DenseNet-121 | DenseNet-169 |
| --- | --- | --- | --- |
| Convolution | 112 x 112 | 7 x 7 Convolution, Stride 2 |  |
| Pooling | 56 x 56 | 3 x 3 Max Pool, Stride 2 |  |
| Dense Block (1) | 56 x 56 | $\begin{bmatrix} 1 \times 1 \text{ conv} \\ 3 \times 3 \text{ conv} \end{bmatrix} \times 6$ | $\begin{bmatrix} 1 \times 1 \text{ conv} \\ 3 \times 3 \text{ conv} \end{bmatrix} \times 6$ |
| Transition Layer (1) | 56 x 56 | 1 x 1 Convolution |  |
|  | 28 x 28 | 3 x 3 Average Pool, Stride 2 |  |
| Dense Block (2) | 28 x 28 | $\begin{bmatrix} 1 \times 1 \text{ conv} \\ 3 \times 3 \text{ conv} \end{bmatrix} \times 12$ | $\begin{bmatrix} 1 \times 1 \text{ conv} \\ 3 \times 3 \text{ conv} \end{bmatrix} \times 12$ |
| Transition Layer (2) | 28 x 28 | 1 x 1 Convolution |  |
|  | 14 x 14 | 3 x 3 Average Pool, Stride 2 |  |
| Dense Block (3) | 14 x 14 | $\begin{bmatrix} 1 \times 1 \text{ conv} \\ 3 \times 3 \text{ conv} \end{bmatrix} \times 24$ | $\begin{bmatrix} 1 \times 1 \text{ conv} \\ 3 \times 3 \text{ conv} \end{bmatrix} \times 32$ |
| Transition Layer (3) | 14 x 14 | 1 x 1 Convolution |  |
|  | 7 x 7 | 3 x 3 Average Pool, Stride 2 |  |
| Dense Block (4) | 7 x 7 | $\begin{bmatrix} 1 \times 1 \text{ conv} \\ 3 \times 3 \text{ conv} \end{bmatrix} \times 16$ | $\begin{bmatrix} 1 \times 1 \text{ conv} \\ 3 \times 3 \text{ conv} \end{bmatrix} \times 32$ |
| Classification Layer | 1 x 1 | 7 x 7 Global Average Pool, Stride 2<br>1000D FC, 2D FC, Softmax |  |

We compared the performance capabilities of the Deep CNNs against traditional Machine learning methodologies, whereby a Principal Component Analysis (PCA)/Support Vector Machine (SVM) based classifier is employed to classify the 24 low-level quantitative vocal descriptors detailed in Section 2.2.1. By calculating the eigenvectors of the covariance matrix of the original inputs, PCA linearly transforms a high-dimensional input vector into a low-dimensional input vector whose components are uncorrelated. The optimal parameters of the Radial Basis Function (RBF) kernel SVM were identified using a grid search approach and C=100 and Gamma = 0.1 exhibited the least classification error.

We trained each DenseNet model for 50 epochs using batch sizes of 8 and applied the Adam optimizer [27] with an initial learning rate of *l*_r_=0.001, which was reduced by a factor of 0.01 when no improvement in validation loss (binary cross-entropy) for five consecutive epochs was observed, to ensure that the model did not overfit during the training process. To prevent group data leakage, we randomly apportioned the data into training, validation and test sets, with an approximate 70*%*-10*%*-20*%* split, ensuring that audio segments originating from the same participant were not contained in both the testing and training data sets. To evaluate the performance of the models in terms of their ability to distinguish between PD and HC participants, we calculated four performance metrics: Accuracy, Sensitivity/Recall, Specificity and the Matthews Correlation Coefficient (MCC) [28].

### 2.4 Saliency Maps

To interpret the network predictions, we generated saliency maps using a gradient-based back-propagation method to visualise which regions of the input mel-spectrogram image influenced the output prediction and reveal which vocal characteristics are most indicative of PD. To highlight class-relevant pixels, the convolutional neural network propagates the output back to the input image space by calculating the gradient of the class score S_class_ with respect to the input pixels, where S_class_ is usually taken to be the activation of the neuron in the output layer encoding the class of interest.

## 3 Results and Discussion

By examining the experimental resultcs presented in Table 3, it can be observed that the DenseNet-169 network architecture conducted on the log-mel-spectrograms of 5-second segmented audio signals achieves the best performance, with a mean sensitivity of 87.9*%* (SD = 5.6*%*) and specificity of 89.4*%* (SD = 7.3*%*). The classification performance of the networks degrade as the length of the audio segment increases, which implies that 10-second segments are perhaps too long to capture subtle vocal microperturbations characteristic of PD. Furthermore, since the sizes of the input log-mel spectrogram images are constant (224×224 pixels), regardless of the duration of the audio segment, segments of a longer duration may lose temporal resolution, thereby reducing their utility for automatic PD discrimination. Learned audio features using deep CNNs marginally outperform the low-level hand-crafted audio features classified with the PCA-SVM model, which suggests that whilst short-term fluctuations in periodic vocal samples of phonatory signals can be quantified using low-level acoustic descriptors (LLD), they may unsuitable for severely disordered voices, which are typically characterised by poor periodicity and from which a period of sustained phonation is harder to extract.

**Table 3.**
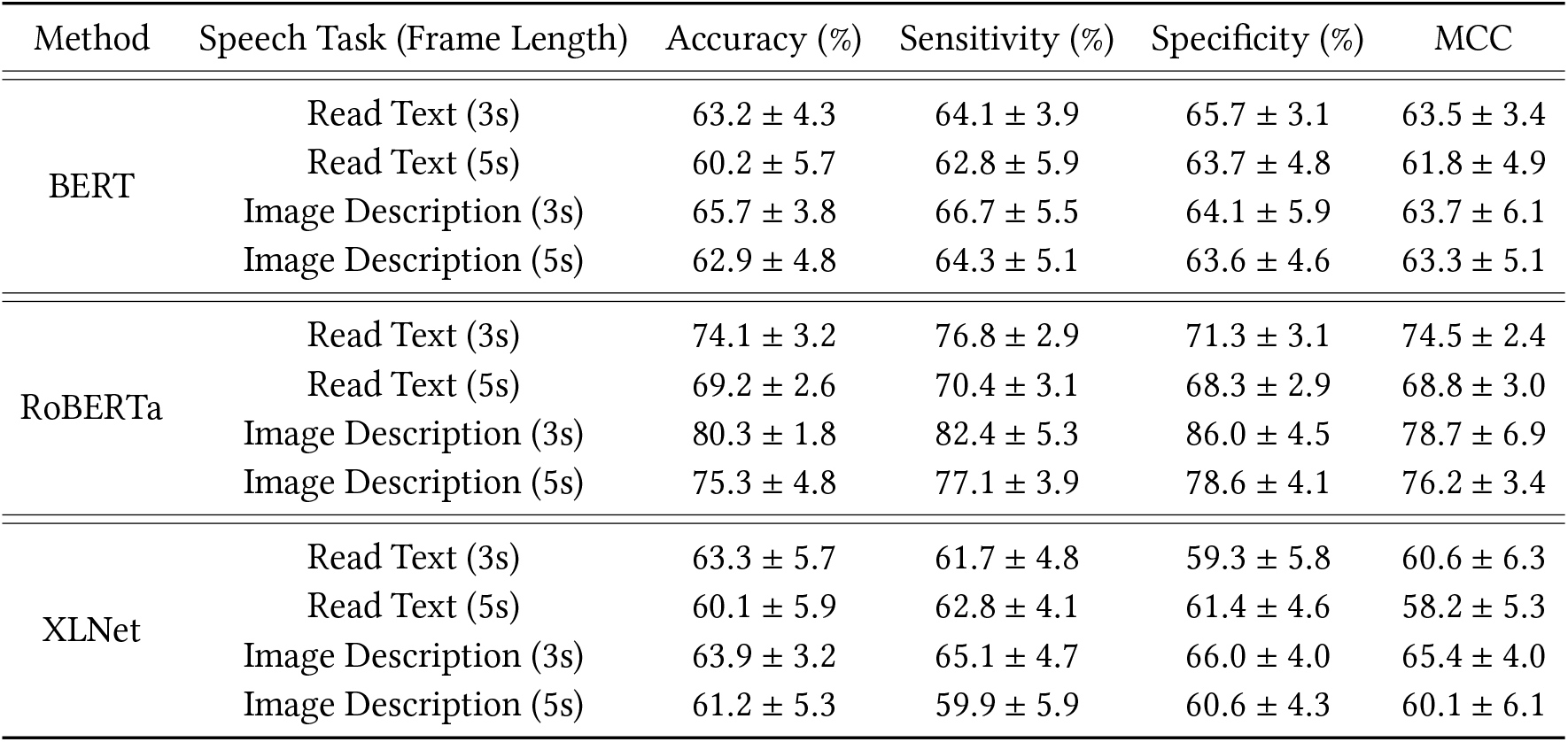
Results for the three large language models trained on ASR-generated transcripts generated using WhisperX (including variable pauses and disfluencies). The results obtained for 10 test runs of each model are presented in the following format: Mean ±Standard Deviation. MCC: Matthews Correlation Coefficient, DN: DenseNet.

| Method | Speech Task (Frame Length) | Accuracy (%) | Sensitivity (%) | Specificity (%) | MCC |
| --- | --- | --- | --- | --- | --- |
| BERT | Read Text (3s) | 63.2 ± 4.3 | 64.1 ± 3.9 | 65.7 ± 3.1 | 63.5 ± 3.4 |
|  | Read Text (5s) | 60.2 ± 5.7 | 62.8 ± 5.9 | 63.7 ± 4.8 | 61.8 ± 4.9 |
|  | Image Description (3s) | 65.7 ± 3.8 | 66.7 ± 5.5 | 64.1 ± 5.9 | 63.7 ± 6.1 |
|  | Image Description (5s) | 62.9 ± 4.8 | 64.3 ± 5.1 | 63.6 ± 4.6 | 63.3 ± 5.1 |
| RoBERTa | Read Text (3s) | 74.1 ± 3.2 | 76.8 ± 2.9 | 71.3 ± 3.1 | 74.5 ± 2.4 |
|  | Read Text (5s) | 69.2 ± 2.6 | 70.4 ± 3.1 | 68.3 ± 2.9 | 68.8 ± 3.0 |
|  | Image Description (3s) | 80.3 ± 1.8 | 82.4 ± 5.3 | 86.0 ± 4.5 | 78.7 ± 6.9 |
|  | Image Description (5s) | 75.3 ± 4.8 | 77.1 ± 3.9 | 78.6 ± 4.1 | 76.2 ± 3.4 |
| XLNet | Read Text (3s) | 63.3 ± 5.7 | 61.7 ± 4.8 | 59.3 ± 5.8 | 60.6 ± 6.3 |
|  | Read Text (5s) | 60.1 ± 5.9 | 62.8 ± 4.1 | 61.4 ± 4.6 | 58.2 ± 5.3 |
|  | Image Description (3s) | 63.9 ± 3.2 | 65.1 ± 4.7 | 66.0 ± 4.0 | 65.4 ± 4.0 |
|  | Image Description (5s) | 61.2 ± 5.3 | 59.9 ± 5.9 | 60.6 ± 4.3 | 60.1 ± 6.1 |

The performance of the DenseNet models diminishes as the depth of the network decreases, thereby demonstrating that an increase in the number of layers leads to an increase in the ability of the network in discriminating between PD and control subjects. The classification performance of the three models is dependent upon the nature of the complex speech-production tasks; although the best results are achieved on the text-dependent running speech task, the results achieved with spontaneous dialogue are not significantly dissimilar which may be because the motor-planning process required to produce spontaneous speech is more complex than monologue tasks and contribute to the appearance of articulation errors and disfluency in the speech of PD patients.

By observing the saliency maps presented Figure 1, it is apparent that the DenseNet-161 classifier is able to distinguish between PD participants and healthy controls by detecting centralised, low-frequency regions of the spectrograms representing the speech of PD subjects, whereas a larger range of frequencies are detected in the spectrograms representing speech from healthy controls. By categorising the PD cohort into two subsets according to the subject’s H&Y score (I–II mild; III–IV severe), it is apparent that there is an equal proportion of participants with early-stage PD (n = 8) as late-stage PD (n=8). The ability of the network in distinguishing between the voices of healthy controls and subjects with early and late-stage Parkinson’s disease is reflected by the final probability distribution generated by the softmax layer for each class, such that high probabilities are associated with late-stage PD (i.e. HY:III-IV) and vice versa.

**Fig. 1.**
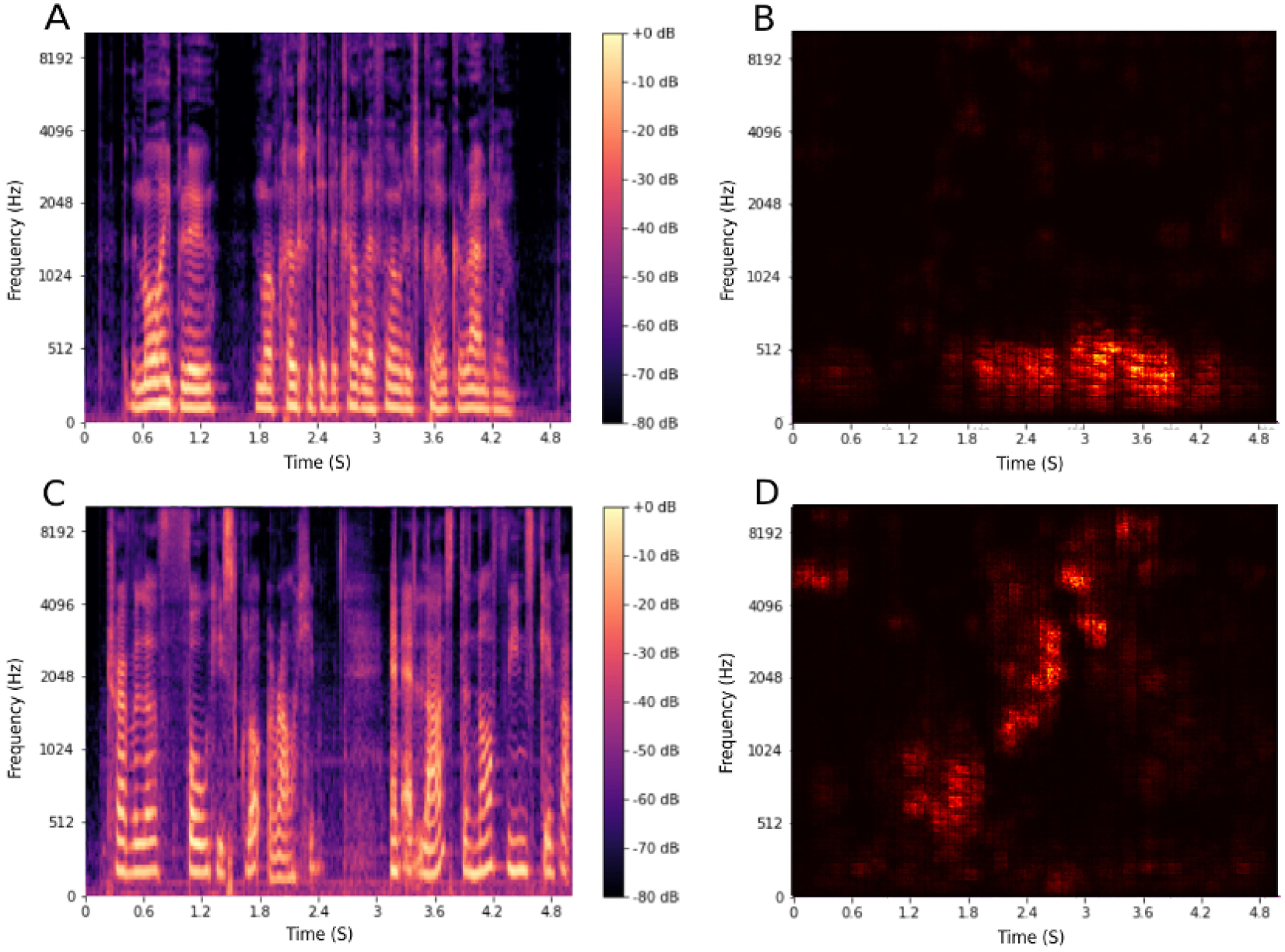
Log mel-spectrogram and saliency map generated by the DenseNet-169 model of a 5s audio segment of the phrase *“*..*immediately the traveller took off his cloak.”* (A) Log Mel-Spectrogram - Parkinson’s Disease (B) Saliency Map - Parkinson’s Disease. (C) Log Mel-Spectrogram - Healthy Control. (D) Saliency Map - Healthy Control.

## 4 Conclusion and Future Works

This work presents an approach that combines audio processing and deep learning techniques in order to develop an assistive tool that can differentiate between the speech of subjects with PD and healthy controls. The limitations of this study include the small sample size, unbalanced gender distribution of subjects and limited additional clinical metadata. Although promising, the findings of this pilot study must be validated using larger, gender-balanced, prospectively- collected datasets, with greater clinical metadata detail, before this technology could be used as a diagnostic decision support tool in a clinical setting.

Future works ought to consider other potential uses of speech and audio analysis techniques for the detection of brain-based pathologies, such as brain tumours or Alzheimer’s, by incorporating the Dense CNN mel-spectrogram classifier proposed in this paper with Natural Language Processing techniques and patient Electronic Health Record (EHR) data in order to develop a multimodal diagnostic tool. Furthermore, the data used in this study was collected in an acoustically controlled environment, therefore these findings could be extended to more realistic acoustic conditions, which would extend the potential of the proposed technology for use in more practical settings.

## Data availability

The Mobile Device Voice Recordings at King’s College London (MDVR-KCL) data set used in this study is publicly available at https://zenodo.org/record/2867216#.YFSECGT7SdZ.

## Credit Authorship Contribution Statement

Study concept and design: S.A.J, H.W, R.K.M. Model development, analysis, and interpretation of results: S.A.J, H.W, R.K.M. Drafting of the manuscript: S.A.J, H.W, R.K.M. Critical revision of the manuscript: S.A.J, J.C, H.W, R.K.M.

## Corresponding Authors

Correspondence to Ryan Mathew or He Wang.

## Conflict of Interest Statement

The authors declare no competing interests.

## Notes

### Competing Interest Statement

The authors have declared no competing interest.

### Funding Statement

This research was supported by the following funding sources: EP/S024336/1 (UKRI Centre for Doctoral Training in Artificial Intelligence for Medical Diagnosis and Care) for Sara Jones; Yorkshire's Brain Tumour Charity 117907 and Candlelighters 117905 for Ryan Mathew.

### Author Declarations

The study used ONLY openly available human data that were originally located at: https://zenodo.org/record/2867216#.YFSECGT7SdZ The Mobile Device Voice Recordings at King's College London (MDVR-KCL) dataset is open-source and publicly accessible through Zenodo. The original study from which this dataset was derived was reviewed and approved by the London - Dulwich Research Ethics Committee (REC reference 17/LO/0909), and all subjects provided written informed consent to participate. As this research involves secondary analysis of openly available data, no separate ethics review is required for our proposed study.

